# Updating reproduction number estimates for mpox in the Democratic Republic of Congo using surveillance data

**DOI:** 10.1101/2023.04.14.23288572

**Authors:** Kelly Charniga, Andrea M. McCollum, Christine M. Hughes, Benjamin Monroe, Joelle Kabamba, Robert Shongo Lushima, Toutou Likafi, Beatrice Nguete, Elisabeth Pukuta, Elisabeth Muyamuna, Jean-Jacques Muyembe Tamfum, Stomy Karhemere, Didine Kaba, Yoshinori Nakazawa

## Abstract

Incidence of human mpox has been increasing in West and Central Africa, including in the Democratic Republic of Congo (DRC), where monkeypox virus (MPXV) is endemic. Most estimates of the pathogen’s transmissibility in DRC are based on data from the 1980s. Amid the global 2022 mpox outbreak, new estimates are needed to characterize the virus’ epidemic potential and inform outbreak control strategies. We used the R package *vimes* to identify clusters of laboratory-confirmed mpox cases in Tshuapa Province, DRC. Cases with both temporal and spatial data were assigned to clusters based on the disease’s serial interval and spatial kernel. We used the size of the clusters to infer the effective reproduction number, *R*_*t*_, and the rate of zoonotic spillover of MPXV into the human population. Out of 1,463 confirmed mpox cases reported in Tshuapa Province between 2013 and 2017, 878 had both date of symptom onset and a location with geographic coordinates. Results include an estimated *R*_*t*_ of 0.82 (95% CI: 0.79 – 0.85) and a rate of 132 (95% CI: 122 – 143) spillovers per year assuming a reporting rate of 0.25. This estimate of *R*_*t*_ is larger compared to most previous estimates. One potential explanation for this result is that *R*_*t*_ could have increased in DRC over time due to declining population-level immunity conferred by smallpox vaccination, which was discontinued around 1982. *R*_*t*_ could be overestimated if our assumption of one spillover event per cluster does not hold. Our results are consistent with increased transmissibility of MPXV in Tshuapa Province.

## Introduction

Mpox (formerly known as monkeypox) is a zoonotic disease that has been endemic in Africa for at least half a century.^1^ The disease is caused by monkeypox virus (MPXV), an orthopoxvirus in the family *Poxviridae*.^2^ The virus is related to variola virus, which causes smallpox,^3^ and is divided into two clades, clade I and clade II. Clade I has historically been found in the Congo Basin, while clade II has historically been found in West Africa.^4^ Vaccination against smallpox using vaccinia virus provides protection against other orthopoxvirus infections; however, routine smallpox immunization ended with the eradication of the disease in 1980. In unvaccinated persons, clade I has an estimated case fatality rate of 11% ^3^ compared to <3% for clade II.^5^ The clinical presentation of mpox is similar to that of smallpox with the exception of the presence of lymphadenopathy in the majority of mpox cases. Classical symptoms begin with a prodrome consisting of fever, headache, muscle aches, fatigue, and lymphadenopathy.^6^ One to three days later, a rash develops, which progresses through several stages over the course of 2 – 4 weeks. Complications from the disease include scarring, permanent corneal scarring leading to loss of vision, bronchopneumonia, encephalitis, and sepsis.^6^

Transmission of MPXV from infected animals to humans occurs via scratches or bites^7^ and may occur while hunting and preparing wild game or contact with infectious fomites (e.g., environmental contamination).^8^ While the animal reservoir(s) is unknown, small mammals including rodents are thought to play a role in the maintenance and spread of the virus.^3^ Following one or more spillover events from the reservoir, human-to-human transmission can occur through close contact with infectious material from skin lesions, respiratory secretions during prolonged face-to-face contact, and fomites such as linens and bedding.^9^ Prior to 2022, human transmission chains as long as seven generations had been observed.^10^

Since 2010, the U.S. Centers for Disease Control and Prevention (CDC) and Kinshasa School of Public Health have been providing support for enhanced surveillance of mpox in Tshuapa Province. Tshuapa Province is located in a rural, forested area of the Congo Basin in the Democratic Republic of Congo (DRC). It has an area of approximately 133,000 km^2^ and a population of approximately 2 million^11^ spread across 12 health zones. The climate is equatorial, with a rainy and a dry season.^12^ The region is characterized by poor infrastructure and widespread poverty.^13^ Much of the population relies on wild game for protein.^14^

A key epidemiological parameter used to understand the transmission potential of infectious diseases is the reproduction number. The reproduction number is defined as the average number of secondary infections generated by a single infected individual.^15^ If the value is above 1, an epidemic is growing, whereas a value below 1 suggests an epidemic is shrinking.^16^ The reproduction number depends on contact patterns, demographic rates, and population-level immunity, among other factors.^17^ If a pathogen is introduced into a population with immunity from vaccination or infection, the reproduction number is considered an effective reproduction number, *R*_t_.^18^ Monitoring *R* for emerging and re-emerging infectious diseases is important for global health security as increases could precede epidemics or pandemics.

There is evidence that incidence of mpox has increased in Central^19, 20, 21^ and West Africa^22^. Most published estimates of *R*_*t*_ for MPXV in DRC are based on data from the 1980s and range from approximately 0.3 – 0.5.^23, 24, 25, 26^ In 2022, a global outbreak of mpox caused more than 85,000 cases in all regions of the world, disproportionately affecting gay, bisexual, and men who have sex with men.^27, 28, 29^ Given the changing epidemiology of mpox, updated estimates of the virus’ transmissibility are needed in historically affected countries. The aim of this study was to identify clusters of mpox cases from surveillance data in Tshuapa Province, DRC and use the cluster sizes to estimate *R*_*t*_ and the rate at which MPXV spills over into the human population.

## Materials and Methods

### Overview

The model framework implemented in the R package *vimes* combines various data types (e.g., temporal, spatial, genetic) to identify clusters of cases in an outbreak. Input data types can include anything for which a pairwise distance can be calculated (e.g., days between symptom onset). *Vimes* connects all cases on a graph for each data type provided. The graph’s edges are weighted by the pairwise distance between cases. Distances greater than specified cutoffs are removed (or “pruned”). The graphs are merged by intersection, with the resulting graph representing clusters of related cases based on all data types. The size of the clusters can then be used to quantify the pathogen’s transmissibility.

### Mpox data

From 2010 – 2019, trained surveillance officers completed paper-based case report forms and collected clinical specimens during investigations of suspected mpox cases in Tshuapa Province (not all suspected cases were investigated). Case report forms include questions about basic demographic information (e.g., age, race/ethnicity, residence), skin lesion characteristics, general signs and symptoms, and exposure history. Lesion swabs, lesion crusts, or blood were tested at the Institut National de Recherche Biomédicale (INRB) in Kinshasa, DRC and CDC in Atlanta, USA Sample processing and laboratory diagnostic methods have been described elsewhere.^30^ Case definitions are provided in Supplementary methods.

We extracted information from line list data on the timing (date of fever or rash onset) and location (village of residence during the last 12 months or village in which rash onset occurred or neighborhood) of infection for laboratory-confirmed mpox cases to identify clusters of disease (Supplementary methods). Cases with missing symptom onset date or missing location were not analyzed.

We calculated the pairwise distance between all cases. For temporal data, the distance was calculated as the difference in days between symptom onset dates of all case pair combinations. For spatial data, the geographic distance between cases was calculated using the Vincenty inverse formula for ellipsoids (implemented using the gdist function in the R package Imap),^31^ given the large area covered by Tshuapa Province.

### Epidemiological parameters

To run *vimes*, we needed one key parameter for each data type: the serial interval distribution for temporal data and the spatial kernel for spatial data. The serial interval distribution is the time between symptom onset in a primary case and symptom onset in a secondary case infected by the primary case.^32^ We used a gamma-distributed serial interval with mean 16.0 days and standard deviation of 3.7 days based on smallpox data^33^ (Supplementary methods). The spatial kernel is the distribution of geographical distances between primary and secondary cases.^32^ We estimated the mean transmission distance of 0.13 km for MPXV using active contact tracing data collected by the World Health Organization (WHO) between 1970 – 1986 (Supplementary methods) and assumed a Rayleigh distribution.

### Cutoffs

Choosing appropriate cutoffs is an important part of using *vimes* as it affects the number of edges kept in the pruning step and the resulting size of clusters.^32^ Cutoffs are informed by prior knowledge on the distribution of distances between cases (i.e., the epidemiological parameters described above). For a particular data type, the cutoff can be defined as a quantile of the input distribution. Two cases are considered unrelated if the distance between them is larger than the cutoff.

### Underreporting

The reporting rate is the percentage of infections that are ultimately reported as suspected or confirmed cases of the disease.^34^ When reporting is low, intermediate cases are missed, and the distance between observed cases can increase. *Vimes* accounts for underreporting by defining the cutoff as *f*^*n, π*^ where *f*^*n*^ is the probability density function or probability mass function of expected distances between a primary and secondary case for data type, n, and n is the probability of a geometric distribution that describes the number of unobserved intermediate cases between two observed cases.

The method assumes surveillance and reporting are stable over the estimation period and that the probability of being reported is the same for all cases. Further details can be found in Cori et al.^32^

Nolen et al. found a reporting rate of 41% in a household study following a 2013 mpox outbreak in Bokungu Health Zone.^10^ We adjusted this reporting rate for partially missing data (cases that were missing symptom onset date or location). We considered the surveillance effort for mpox in Tshuapa Province to be relatively stable from 2013 – 2017, based on the total number of suspected mpox cases investigated and the percentage of investigated cases that were confirmed by year and health zone

### Estimation of *R*_*t*_ and spillover rate

We estimated *R*_*t*_ and the rate of spillover of MPXV using the R package *branchr*.^35^ The method assumes one zoonotic spillover event per cluster. Further details can be found in Supplementary methods.

### Sensitivity analyses

We repeated the analysis by year and by health zone as sensitivity analyses. We also examined the impact of the cutoffs (quantiles) and assumption regarding reporting rates on our estimates, including the use of a shorter serial interval (Supplementary methods).

### Simulations

As recommended by Cori et al.,^32^ we performed simulations to find the optimal cutoffs for the specific context of mpox in Tshuapa Province and check that the method can correctly identify mpox clusters as well as accurately estimate *R*_*t*_ and the spillover rate.

### Validation

We searched the Program for Monitoring Emerging Diseases (ProMED) and Google Scholar for documentation of any outbreaks of mpox in Tshuapa Province that may have occurred during the study period. To further evaluate the plausibility of identified clusters, we compared the calculated pairwise differences by data type and cluster^36^ for singletons and clusters with 10 or more cases. We would expect clusters to have smaller pairwise distances compared to the distribution of pairwise distances for singletons.

We checked the assumption of one spillover event per cluster using exposure history. If reporting were perfect and our assumption were correct, we would expect that patients assigned to larger clusters would report contact with an ill person prior to symptom onset more frequently than singletons (cases not linked to any other cases).

### Ethics statement

Surveillance was conducted in agreement with Congolese national guidelines. The activity was determined to not be research by a CDC human subjects advisor (project determination RD-071811MR).

### Availability

All analyses were conducted in R version 4.1.1. Simulation code is available on GitHub (https://github.com/kcharniga/mpox_in_drc). The mpox line list and geographic data associated with cases are owned by the DRC Ministry of Health. The decision to release these data rests with the Ministry of Health.

## Results

### Mpox surveillance and reporting

From 2010 – 2019, a total of 2,993 suspected cases were investigated for mpox in Tshuapa Province (Table 1). Of the investigated cases, 43 (1%) were excluded from the analysis due to inconclusive or missing laboratory results, 2,019 (67%) tested positive for *Orthopoxvirus* or MPXV, and 931 (31%) tested negative for *Orthopoxvirus* or MPXV. On average, 299 (range 11 – 526) suspected cases were investigated for mpox in Tshuapa Province each year (Table 1). The number of investigated mpox cases increased nonmonotonically from 2010 – 2016 and declined each year after 2016. The percentage of investigated mpox cases that were confirmed was low for the first three years (mean 36%) before stabilizing for the last seven years (mean 76%, range 70% – 80%).

**Table 1.**
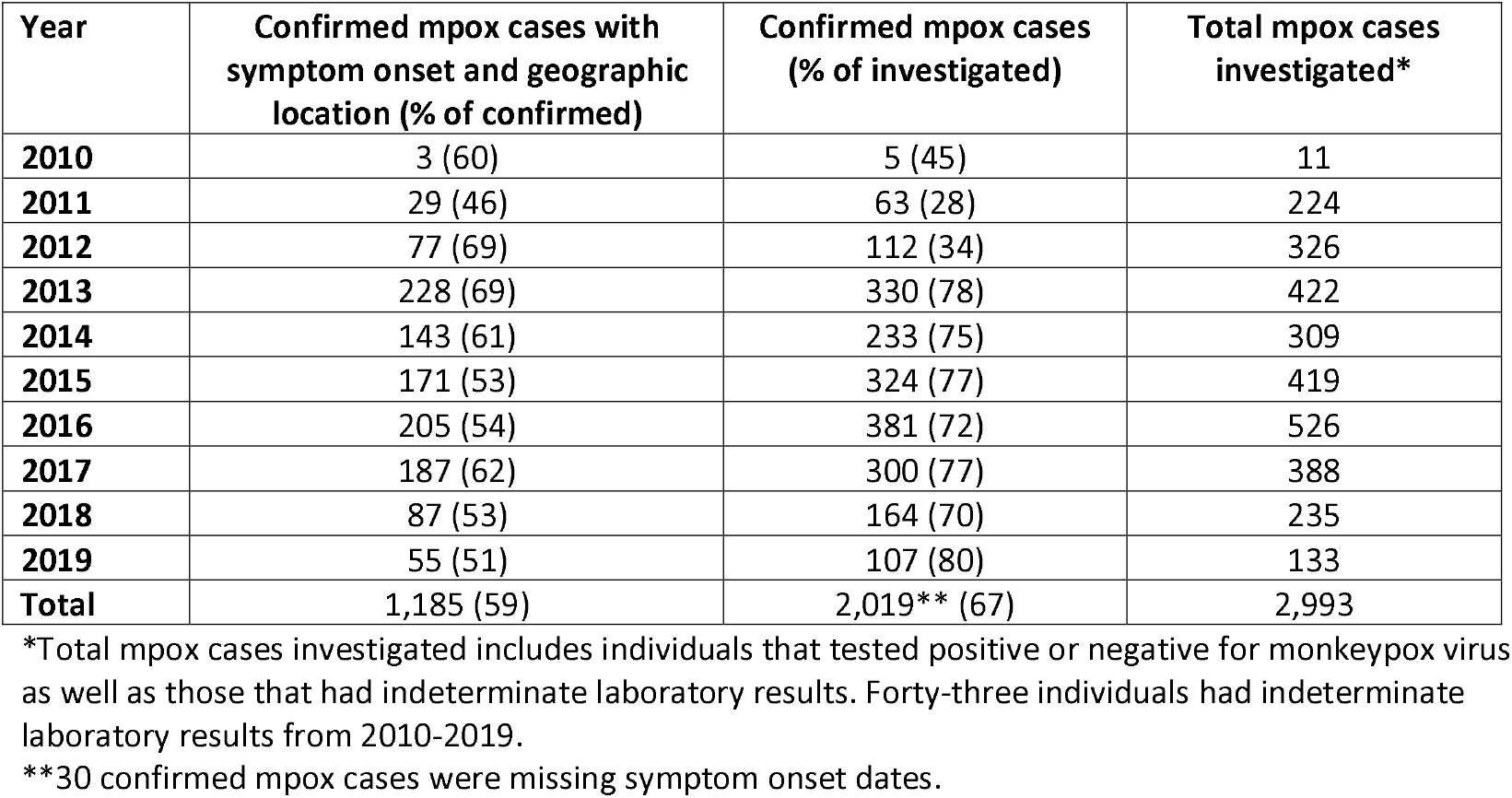
Human mpox reporting by year from surveillance in Tshuapa Province, DRC, 2010 – 2019.

| Year | Confirmed mpox cases with symptom onset and geographic location (% of confirmed) | Confirmed mpox cases (% of investigated) | Total mpox cases investigated* |
| --- | --- | --- | --- |
| 2010 | 3 (60) | 5 (45) | 11 |
| 2011 | 29 (46) | 63 (28) | 224 |
| 2012 | 77 (69) | 112 (34) | 326 |
| 2013 | 228 (69) | 330 (78) | 422 |
| 2014 | 143 (61) | 233 (75) | 309 |
| 2015 | 171 (53) | 324 (77) | 419 |
| 2016 | 205 (54) | 381 (72) | 526 |
| 2017 | 187 (62) | 300 (77) | 388 |
| 2018 | 87 (53) | 164 (70) | 235 |
| 2019 | 55 (51) | 107 (80) | 133 |
| **Total** | **1,185 (59)** | **2,019\*\* (67)** | **2,993** |

\*Total mpox cases investigated includes individuals that tested positive or negative for monkeypox virus as well as those that had indeterminate laboratory results. Forty-three individuals had indeterminate laboratory results from 2010-2019.
\*\*30 confirmed mpox cases were missing symptom onset dates.

During the period of stable reporting from 2013 – 2017, there were 1,568 laboratory-confirmed cases of mpox reported in Tshuapa Province (Table 2). Of those, 934 (60%) had complete information on symptom onset and geographic location. There were 477 male cases (51%) out of 932 with available data. The mean age of cases was 16 (range 0 – 79) years (12 cases were missing age). During this period, the number of suspected mpox cases that were investigated ranged from 40 in Monkoto to 300 in Djolu (Table 2). The percentage of investigated cases that were confirmed ranged from 65% in Mondombe to 90% in Wema, and the percentage of confirmed mpox cases with complete temporal and spatial information ranged from 41% in Ikela to 81% in Djolu.

**Table 2.** Human mpox reporting by health zone from surveillance in Tshuapa Province, DRC, 2013 – 2017.

| Health zone | Confirmed mpox cases with symptom onset and geographic location (% of confirmed) | Confirmed mpox cases (% of investigated) | Total investigated | Estimated population size* (2016) |
| --- | --- | --- | --- | --- |
| **Befale** | 32 (44) | 72 (73) | 99 | 177,864 |
| **Boende** | 59 (53) | 111 (70) | 158 | 272,699 |
| **Bokungu** | 80 (54) | 147 (74) | 198 | 201,658 |
| **Busanga** | 152 (73) | 208 (76) | 272 | 100,372 |
| **Djolu** | 166 (81) | 206 (69) | 300 | 235,873 |
| **Ikela** | 60 (41) | 148 (77) | 191 | 198,471 |
| **Lingomo** | 99 (55) | 179 (85) | 210 | 135,617 |
| **Mompono** | 106 (78) | 136 (85) | 160 | 142,956 |
| **Mondombe** | 59 (48) | 122 (65) | 189 | 173,630 |
| **Monkoto** | 16 (59) | 27 (68) | 40 | 139,186 |
| **Wema** | 39 (42) | 92 (90) | 102 | 114,787 |
| **Yalifafu** | 66 (55) | 120 (83) | 145 | 174,973 |
| **Total** | 934 (60) | 1,568 (76) | 2,064 | 2,068,084 |

\*We used population data for Tshuapa Province from 2015 (Annuaire Statistique 2015, Institut National de la Statistique, DRC) and applied these to demographic estimates for each health zone (unpublished, Division Provinciale de la Santé, Tshuapa). Data for 2016 were extrapolated assuming an annual growth of 3.3%.

### Cluster identification and parameter estimation

We identified 334 clusters, 161 (48%) of which were singletons (Figure 1 and Figure S2). The largest cluster size was 30, and the mean cluster size was 2.8. We estimated an *R*_*t*_ of 0.82 (95% confidence interval [CI]: 0.79 – 0.85) and an annual spillover rate of MPXV into the human population of 132 (95% CI: 122 – 143) (Table S1). We obtained these results assuming a reporting rate of 25% (41% from Nolen et al.^10^ times 60% of confirmed cases with complete data) and a cutoff corresponding to the 95%^1/3^ (or 98.3%) quantile of the input distributions. These cutoffs correspond to 233 days and 0.7 km.

**Figure 1.**
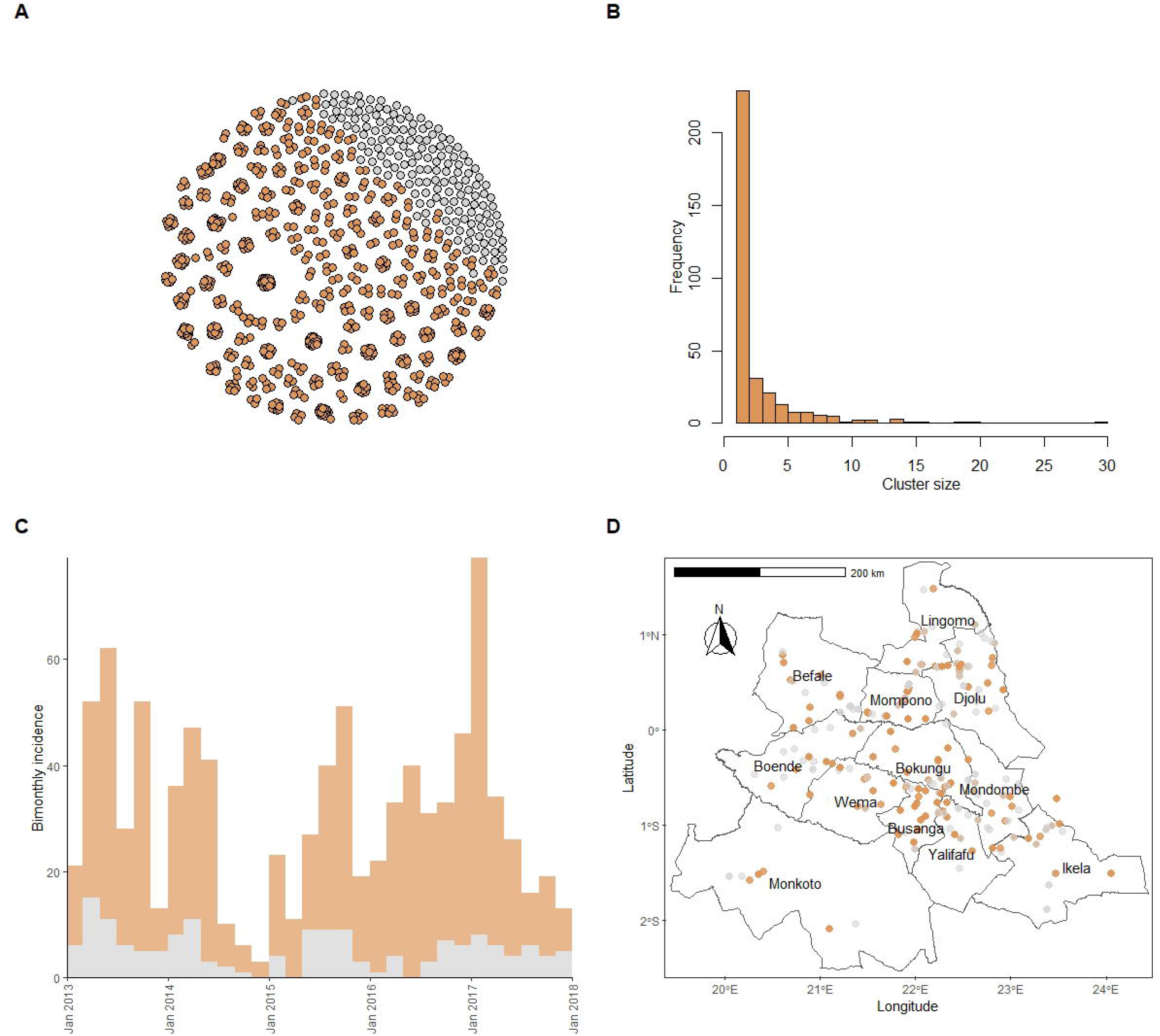
Clusters of human mpox cases in Tshuapa Province, DRC, 2013 - 2017. Nodes represent cases, while edges represent possible epidemiological links. (B) Frequency of cluster sizes. (C) Bimonthly incidence of human mpox cases by date of symptom onset. Mpox reporting tends to decline in December/January each year, likely due to the holidays and healthcare worker trainings. (D) Map of Tshuapa Province showing the geographic locations of human mpox cases. We adjusted the transparency of the points to improve visualization but overplotting still occurs for some villages. We assumed a reporting rate of 25% and used pruning cutoff distances associated with the 98.3% quantiles of the input distance distributions. For (A), (C), and (D), cases in orange belong to clusters containing two or more cases, while cases in gray are not connected to other cases, using both temporal and spatial data. For (D), we downloaded shapefiles of health zones in DRC from the Humanitarian Data Exchange. The shapefiles were prepared by the American Red Cross and are available under an Open Database License (ODC-ODbL).

### Sensitivity analyses

As expected, using higher cutoffs led to fewer, larger clusters in sensitivity analyses. Using cutoffs ranging from the 90% and 98.3% quantiles of the input distributions, the mean cluster sizes ranged from 2.2 – 3.6, and the maximum cluster size was stable (between 29 and 30). Other summary statistics can be found in Table S5. Higher cutoffs also led to higher estimates of both *R*_*t*_ and spillover rate (Figure S6). The estimated *R*_*t*_ for the most extreme combinations considered (90% quantile/50% reporting rate and 98.3% quantile/10% reporting rate) was 0.67 (95% CI: 0.63 – 0.71) and 0.92 (95% CI: 0.90 – 0.94) respectively, and the estimated annual spillover rate was 123 (95% CI: 116 – 130) and 151 (95% CI: 136 – 168) respectively. Using cutoffs corresponding to a shorter serial interval estimated during the global mpox outbreak did not have a major impact on results; however, setting the distance cutoff to 10 km increased the estimated *R*_*t*_ to 0.89 (95% CI: 0.86 – 0.93) (Supplementary results, Figure S8, and Table S7). Results of the analysis by year and health zone can be found in Supplementary results (Figures S10 – S13, Tables S2 – S3). While *R*_*t*_ was not different across years or health zone (the CIs overlapped), there was heterogeneity in the annual spillover rate by year and health zone (some CIs did not overlap).

### Simulations

Our simulation study showed that the optimal cutoff for mpox in Tshuapa Province is defined by the 98.3% quantile (Figure S15). Further simulation study results can be found in Supplementary results and Figure S16.

### Validation

We found two reports of potential mpox outbreaks in Tshuapa Province during the study period. Details can be found in Supplementary results.

The calculated pairwise differences by data type and cluster for singletons and clusters with 10 or more cases are shown in Figure S17. As expected, we found that the median pairwise differences were significantly lower on the temporal and spatial dimensions in all clusters compared to those dimensions in the singletons. In Figure S18, we present the epidemiologic curves associated with these clusters. While cases occur sporadically over the time series for some clusters, others have uninterrupted incidence.

We evaluated exposure information for the 934 mpox patients included in our cluster analysis to check the assumption of one spillover event per cluster. Nearly half of patients with available data reported contact with a person or persons presenting with similar symptoms in the three weeks prior to symptom onset, nearly half reported having touched a wild animal during three weeks prior to symptom onset, and 13% reported both types of contact (Supplementary results). The distribution of patients who reported contact with ill people or animals was similar regardless of cluster type (singleton vs cluster with size > 1, Figure S3). We found some statistically significant differences in the proportion of mpox patients who reported contact with ill people or animals by health zone (Figure S4).

## Discussion

We found that *R*_*t*_ for MPXV in rural DRC has increased compared to previous decades and is consistent with estimates from a study published in 2020.^37^ Increases in *R* t since the 1980s^23, 24, 25, 26^ could be attributed to declining population-level immunity conferred by smallpox vaccination, behavior change, including increased movement of people, ecological and environmental changes, and/or changes that may predispose the virus to spread more easily between humans. Spatial heterogeneity in the annual spillover rate could be explained by different levels of exposure to potential animal reservoirs of MPXV across health zones.

We assumed surveillance of mpox was relatively stable from 2013 – 2017 based on the total number of suspected cases that were investigated. However, an Ebola outbreak in Tshuapa Province may have affected mpox reporting in the second half of 2014, when 69 cases of Ebola virus disease were reported in Boende Health Zone near the border with Wema.^38^ Consequently, all specimen collection for non-Ebola virus disease in Tshuapa Province was halted for at least six months. Also, from 2012 – 2015, epidemiological studies of mpox were conducted in Djolu, Busanga, and Bokungu, which may have increased the rate of case investigations.

The cutoff we used for spatial distance (0.7 km) is conservative, and the mean transmission distance informing this cutoff is likely underestimated. We assumed within-village transmission in the WHO data occurred over a distance of 0 km; if case pairs belonged to different households, the pairwise distance would be non-zero. Also, we were not able to find geographic coordinates for the villages of six case pairs with mismatching villages. Although the estimated transmission distance is based on data collected in the 1970s and 80s, the transportation infrastructure in the region has not changed substantially since then.

The serial interval estimate we used to inform the temporal cutoff agreed with the historical range observed for MPXV in DRC^39^ but ultimately was based on data for variola virus.^33^ The serial interval has been estimated for MPXV using data from the 2022 global outbreak by several groups.^40, 41, 42^ For example, Madewell et al. reported an estimated mean serial interval of 8.5 days (95% credible interval [CrI]: 7.3 – 9.9) in the U.S.^40^ Slightly longer serial intervals were reported from Europe, with mean 9.5 days (95% CrI: 7.4 – 12.3) reported in the U.K.^42^ and mean 10.1 days (95% CI: 6.6 – 14.7) reported in the Netherlands.^41^ However, these estimates could differ from those in rural DRC due to viral genetic differences, including different clades, and the route and intensity of exposure. According to data from the 2003 mpox outbreak in the U.S., which was linked to the exotic pet trade, people who were exposed to MPXV by non-invasive routes, such as petting an infected animal, experienced slower illness progression and longer incubation period than those with complex exposures, such as a bite or scratch from an infected animal.^7^ Because the serial interval and incubation periods are correlated, we would expect longer incubation periods (and therefore, longer serial intervals) in rural DRC, where human-to-human transmission seems driven by household exposures,^10^ compared to the global outbreak. Nevertheless, using an estimated serial interval from the global outbreak did not have a large impact on results.

The gold standard for validating clusters of disease is an epidemiological link between cases.^36^ Although some mpox case report forms in our study list contacts, identifying those contacts is challenging. The identification of mpox clusters in our study could be improved with more available data types, such as social, ecological, and genetic data. *Vimes* is flexible in that it can use any measure of distances between cases. Genetic data were only available for a small proportion of mpox cases in Tshuapa Province.

We did not find published reports about several large clusters of mpox cases identified in our study, including one consisting of 30 patients from the village of Mbotolongo in Djolu in 2017. While outbreaks of Ebola, a viral hemorrhagic fever disease, in DRC often garner international media coverage^43, 44^ and resources,^45^ outbreaks of other diseases, such as mpox,^46^ measles,^47^ and yellow fever,^48, 49^ receive less attention. At the same time, we were not able to capture at least one large outbreak (consisting of 13 cases from October – November 2016) in our study because the geographic coordinates of the village (Bowe in Wema Health Zone) were unknown.

The high proportion of confirmed mpox cases in Tshuapa Province with missing location data highlights the need to collect better geographic data in the country. These data could be used to improve other public health programs, such as routine childhood immunizations.^50^ Although not required for this analysis, demographic data for DRC are limited. Current population estimates are projections based on the last national census, which was conducted nearly four decades ago.^51^

One limitation of our study is that we used a reporting rate estimated from one health zone at one time point.^10^ We do not know to what extent the reporting rate varied between health zones or across years during the study period. We do know that reporting was low, especially at the national level. Hoff et al. estimated that suspected mpox cases in DRC were 5 – 15 times higher than what was reported (i.e., reporting rates of 7% – 20%) in 2013.^52^

An additional limitation is that fever may not be accurately recalled by cases^10^ as several other febrile diseases are common in DRC. Alternatively, we could have used date of rash onset in our study but wanted to capture the earliest date of symptom onset, which is usually marked by fever. Another limitation is the timeliness of the data. Data for 2020 – 2022 were incomplete at the time of writing. Delays can be attributed to lack of equipment and infrastructure, the need for confirmatory testing at CDC, and the use of paper forms for data collection, among others. Finally, using exposure information to check the assumption of one spillover event per cluster was complicated by large underreporting and the fact that many people are exposed to both sick people and animals in and around their homes. *R*_*t*_ could be overestimated if this assumption does not hold. Future studies should continue to characterize the relationship between humans and potential animal reservoirs of MPXV in Central Africa.

## Data Availability

Simulation code is available on GitHub (https://github.com/kcharniga/mpox_in_drc). The mpox line list and geographic data associated with cases are owned by the DRC Ministry of Health. The decision to release these data rests with the Ministry of Health.

## Acknowledgements

We thank all the medical and public health professionals involved in investigating and reporting mpox cases in Tshuapa Province. Thanks to Pierre Nouvellet for helpful comments on the manuscript.

## Disclosures

We declare no conflicts of interest.

## Disclaimer

The findings and conclusions in this report are those of the authors and do not necessarily represent the official position of the Centers for Disease Control and Prevention, U.S. Department of Health and Human Services.

## Author contributions

Conceptualization (KC, AMM, YN), methodology (KC), software (KC), validation (KC), formal analysis (KC), investigation (KC), data curation (all authors), writing – original draft (KC), writing – review & editing (all authors), visualization (KC), supervision (YN, AMM).

## Notes

### Competing Interest Statement

The authors have declared no competing interest.

### Funding Statement

This study did not receive any funding.

### Author Declarations

Surveillance was conducted in agreement with Congolese national guidelines. The activity was determined to not be research by a Centers for Disease Control and Prevention human subjects advisor (project determination RD-071811MR).

