## Appendix for "Updating reproduction number estimates for mpox in the Democratic Republic of Congo using surveillance data"

**Supplementary methods**

**Supplementary results**

**Supplementary figures**

**Figure S1.** Distributions of the serial interval (top) and distance kernel (bottom) used to identify clusters of human mpox cases in Tshuapa Province, DRC.

**Figure S2.** Clusters of mpox cases in Tshuapa Province, DRC, 2013 – 2017 obtained using (A) only temporal distances, (B) only spatial distances, and (C) both data types combined.

**Figure S3.** MPXV exposure history by cluster type.

**Figure S4.** MPXV exposure history by health zone.

**Figure S5.** Distributions of pairwise distances for reported human mpox cases in Tshuapa Province, DRC.

**Figure S6.** Estimated effective reproduction number, *R_t_*, (top) and annual rate of spillover of monkeypox virus into the human population (bottom) in Tshuapa Province, DRC, 2013 – 2017 for different assumptions about the reporting rate (10%, 25%, or 50%) and cutoff for pruning (90%, 95%, or 98.3% quantile).

**Figure S7.** Violin plots of the distribution of outbreak length in days in Tshuapa Province, DRC, 2013 – 2017 for different assumptions about the reporting rate (10%, 25%, or 50%) and cutoff for pruning (90%, 95%, or 98.3% quantile).

**Figure S8.** Estimated effective reproduction number, *R_t_*, (top) and annual rate of spillover of monkeypox virus into the human population (bottom) in Tshuapa Province, DRC, 2013 – 2017 for different assumptions about the reporting rate (10%, 25%, or 50%) and cutoff for pruning (90%, 95%, or 98.3% quantile) using a serial interval with mean 10.3 days.

**Figure S9.** Violin plots of the distribution of outbreak length in days in Tshuapa Province, DRC, 2013 – 2017 for different assumptions about the reporting rate (10%, 25%, or 50%) and cutoff for pruning (90%, 95%, or 98.3% quantile) using a serial interval with mean 10.3 days.

**Figure S10.** Clusters of human mpox cases in Tshuapa Province, DRC by year 2013 – 2017.

**Figure S11.** Estimated effective reproduction number, *R_t_*, (top) and annual rate of spillover of monkeypox virus into the human population (bottom) in Tshuapa Province, DRC by year 2013 – 2017.

**Figure S12.** Clusters of human mpox cases in Tshuapa Province, DRC by health zone, 2013 – 2017.

**Figure S13.** Estimated effective reproduction number, Rt, (top) and annual rate of spillover of monkeypox virus into the human population (bottom) in Tshuapa Province, DRC by health zone, 2013 – 2017.

**Figure S14.** Distribution of the simulated dataset sizes for each of the four simulation scenarios considered.

**Figure S15.** Distribution of the optimal cutoff choice for mpox in Tshuapa Province, DRC.

**Figure S16.** The model performance for the control simulation using different reconstruction scenarios is shown in (A) and (B). The model performance for the baseline reconstruction scenario (using the optimal cutoff of 98.3%) applied to simulation scenarios with different reporting rates (low, high, and perfect) is shown in (C) and (D).

**Figure S17.** Notched boxplots of the pairwise differences by data type and cluster of human mpox cases, Tshuapa Province, DRC, 2013 – 2017.

**Figure S18.** Weekly incidence of human mpox cases by date of symptom onset for clusters with at least 10 cases, Tshuapa Province, DRC, 2013 – 2017.

**Supplementary tables**

**Table S1.** Published estimates of the basic reproduction number (*R_o_*) and effective reproduction number (*R_t_*) for mpox in DRC.

**Table S2.** Characteristics of clusters of human mpox cases in Tshuapa Province, DRC by year, 2013 – 2017. Clusters were identified using an algorithm implemented in the R package *vimes*.

**Table S3.** Characteristics of clusters of human mpox cases in Tshuapa Province, DRC by health zone, 2013 – 2017.

**Table S4.** Cutoffs for pairwise distances considered in sensitivity analyses.

**Table S5.** Sensitivity analysis on cutoffs and reporting rates for identifying clusters of human mpox cases in Tshuapa Province, 2013 – 2017.

**Table S6.** Cutoffs for pairwise distances considered in sensitivity analyses using a serial interval with mean 10.3 days.

**Table S7.** Sensitivity analysis on quantiles and reporting rates for identifying clusters of human mpox cases in Tshuapa Province, 2013 – 2017 using a serial interval with mean 10.3 days.

**Table S8.** Simulation scenarios.

**Table S9.** Reconstruction scenarios.

**References**

**Supplementary methods**

*Mpox case definitions*. We defined a suspected mpox case as a person with a vesicular or pustular rash with deep-seated, hard pustules and at least one of the following symptoms: fever preceding the eruption, lymphadenopathy (inguinal, axillary, or cervical), or pustules or crusts on the palms of the hands or soles of the feet. A confirmed mpox case needed to have at least one clinical specimen that tested positive for *Orthopoxvirus* or MPXV DNA with real-time polymerase chain reaction (PCR).

*Identifying clusters*. For the time variable, we used the date of fever onset. If the date of fever onset was missing, we used the date of rash onset instead. We used date of fever onset because fever typically occurs before rash during mpox illness, and the onset of symptoms usually coincides with the infectious period.^1^ For location, we used the village of residence during the last 12 months. If this location was missing, we used the village in which rash onset occurred. If both these variables were missing, we used quartier (neighborhood). If more than one village was listed on the case report form separated by a slash (/), we used the first one, assuming either the patient spent more time there or that location was more relevant for their infection.

*Unique localities dataset.* We compiled a dataset of unique localities in DRC (N = 461) consisting of village name, health zone, and geographic coordinates. Geographic coordinates were compiled from a variety of sources, including GPS, landcover maps,^2^ the Humanitarian Data Exchange,^3, 4^ Map for Environment,^5^ Joint Operation Graphics (JOG) topographic reference maps,^2^ the National Geospatial-Intelligence Agency’s (NGA) Geographic Names Server (GNS) database,^6^ Google Earth,^7^ and hand-drawn maps compiled with input from local healthcare workers. GPS data were collected by collaborators in the field; we considered these data to be the most reliable, while georeferenced hand-drawn maps were the least reliable. We cross-checked village locations with multiple data sources whenever possible. Villages were excluded from the dataset if they shared a name with at least one other village in the same health zone. Villages with the same name that were numbered, i.e., Yolonga 1, Yolonga 2, etc., within a health zone were considered unique and included.

*Cleaning geographic data*. We obtained the geographic coordinates for cases by matching the village name and health zone from the line list with the corresponding information in the unique localities dataset. For combinations of villages and health zones without an exact match, we followed up with fuzzy matching using the fuzzyjoin package in R (version 0.1.6). We used the Jaro method (“jw”, p = 0) which is recommended for possible typos resulting from human-typed text strings.^8^ We performed a second round of matching (exact followed by fuzzy) for cases that still did not have a match that had different villages listed for village of residence during the last 12 months and village in which rash onset occurred.

There are several potential sources of error for the location data, and hence, reasons why a village was classified as “missing” for a patient. A common reason was that nothing was written on the case report form for either residence or village of rash onset. In some instances, a village name was written on the form, but the handwriting was illegible. Village names could have been misspelled on the case report forms, or they could have been spelled correctly on the forms but entered incorrectly into the database (we addressed the latter issue by double checking the case report forms). Even small misspellings can cause doubts about the location of a case. For example, 44 km separates the villages of Lofondo and Lofonda in Befale Health Zone. Villages were also considered missing if they were not unique. A village was not unique if it shared a name with another village in the same health zone or if we did not have geographic coordinates for it.

*Serial interval estimation*. The serial interval depends on the incubation period (the time from infection to symptom onset), the infectivity of the primary case, and population contact patterns. We assumed the serial interval for mpox would be similar to smallpox and fitted a gamma distribution to observed serial interval counts for smallpox (we digitized Fig. 2b^9^ using WebPlotDigitizer^10^). We obtained a shape parameter of 18.5 and a rate parameter of 1.2. These values correspond to a mean serial interval of 16.0 days with a standard deviation of 3.7 days which agrees with the range of the serial interval for MPXV observed in DRC (7 – 23 days, Figure S1).^11^

*Mean transmission distance.* We estimated the mean transmission distance of MPXV from mpox surveillance data collected by the World Health Organization (WHO) between 1970 – 1986. Active contact tracing efforts were initiated in seven Central and West African countries: Cameroon, Central African Republic, Ivory Coast, Liberia, Nigeria, Sierra Leone, and Zaire (present-day Democratic Republic of Congo or DRC), during the smallpox eradication program.^2^

Overall, 404 cases of human mpox were confirmed, with 386 from DRC. One-hundred and forty-four mpox case pairs were identified in DRC;^12^ of those, 92 case pairs were reported in Equateur Region, which includes present-day Tshuapa Province. The contact tracing data did not distinguish between secondary transmission and co-primary infections. As we were only interested in the former, we removed case pairs that had fever onset (or rash onset if fever onset was missing) less than 7 days or more than 23 days apart, the historical range of the serial interval for person-to-person spread of mpox.^11^ Sixty case pairs remained; of those, 53 had the same locality listed and 7 had mismatched localities. For case pairs with matching localities, we assumed the distance over which transmission occurred was 0 km. Out of 7 case pairs with mismatched localities, we could only find the geographic coordinates for the two localities of one case pair. The localities were Yayama and Yaisese, both in Ikela Health Zone. We calculated a geodesic distance between them of 7 km using the Vincenty inverse formula for ellipsoids using the gdist function in the *Imap* package (version 1.32) in R.^13^ We dropped the 6 case pairs with unknown locations.

The mean transmission distance for person-to-person spread of MPXV in Equateur region was calculated as $\frac{0 km \times53 case pairs + 7 km \times1 case pair}{54}= 0.13 km$, which corresponds to a Rayleigh distributed spatial kernel with scale 0.10 km (Figure S1).

*Estimation of R_t_ and spillover rate.* The method implemented in the R package *branchr* to estimate *R_t_* assumes that the number of secondary cases follows a Poisson distribution with mean *R_t_*. The likelihood of the observed final size of the outbreak is adjusted by integrating over all possible unobserved cases. This approach assumes that unobserved cases are distributed randomly across the outbreaks.^14^ The probability of observing an outbreak, $P_{obs}$, is also estimated. The number of unobserved outbreaks is estimated by assuming a negative binomial distribution with parameters *n* and $P_{obs}$, for a given *R_t_* and reporting rate. The total number of spillovers is the estimated number of observed outbreaks (clusters) plus the estimated number of unobserved outbreaks. A rate is obtained by dividing this result by the time over which surveillance occurred.^14^

*Sensitivity analyses on cutoffs.* The results presented in the main text correspond to a reporting rate of 25% and a cutoff corresponding to the 98.3% quantile of the input distance distributions. However, we considered nine total combinations of quantiles (90%, 95%, and 98.3%) and reporting rates (10%, 25%, and 50%) (Figure S5). The cutoffs correspond to distances between cases ranging from 62 – 595 days and from 0.3 – 1.1 km (Table S4). Lower reporting and higher quantiles are associated with larger cutoffs.

We also performed this sensitivity analysis (nine combinations of quantiles and reporting rates) using a serial interval estimated from the global mpox outbreak. Miura et al. reported a gamma-distributed serial interval with mean 10.3 days and standard deviation 6.3 days from 34 case pairs in the Netherlands.^15^ The cutoffs can be found in Table S6 and range from 43 – 404 days.

An additional sensitivity analysis involved manually setting the spatial distance cutoff at 10 km to account for potential underestimation of the spatial kernel (we used the temporal cutoff corresponding to that used in the main results, 233 days).

We used the cluster sizes resulting from these 19 combinations of quantiles and reporting rates to estimate *R_t_* and the annual spillover rate of MPXV.

*Simulations*. We examined four simulation scenarios representing the transmission of MPXV among humans (Table S8). The baseline scenario was meant to resemble the transmission dynamics associated with mpox in our surveillance dataset from Tshuapa Province. For the other three scenarios, we used the same simulated datasets but varied the reporting rate.

For each simulation scenario, we used *vimes* to reconstruct the clusters of mpox cases and re-estimate both *R_t_* and the spillover rate. We used the same input distributions as in the simulations for the serial interval and spatial kernel. For the baseline simulation scenario, we varied the cutoffs and reporting rate (Table S9).

Following Cori et al.,^14^ we adapted an existing simulation implemented in the simOutbreak function of the outbreaker^16^ R package and utilized helper functions in the R package quicksim (version 0.0.1).^17^

The simulation uses a simple branching process. We used a rectangle of 446 km x 413 km as the geographic area for the simulation. The bounds for the rectangle were chosen by taking the distance between mpox cases located in villages with the largest and smallest longitudes and latitudes for the length (x direction) and width (y direction), respectively. The distance between the coordinates was calculated using the Vincenty inverse formula for ellipsoids (implemented using the gdist function in the R package Imap).^13^ We initialized each simulation with a single mpox case placed randomly in the rectangle with symptom onset on day 0. For each day *t* after day 0, both local transmission and spillover from the animal reservoir led to new MPXV infections.

We drew the number of newly infected mpox cases arising from local transmission from a Poisson distribution with mean $\lambda= R_{0}\sum_{s=1}^{t} I_{t-s}w_{s}$. In this equation, *R_0_* is the basic reproduction number, *I_t-s_* is the total number of cases that occurred at time *t-s* (through both local transmission and spillover), and *w* is the probability mass function of the serial interval. *R_0_* is the average number of secondary infections generated by a single infected individual in a large, completely susceptible population.^18^

The next step was to assign an infector to each newly infected case among cases that already occurred. To do this, we used weights for cases that occurred at time *t-s* equal to *w_s_*. In other words, we chose the infectors for each new case according to their current infectivity. The location in the rectangle (x and y coordinates) of each newly infected case came from drawing an *x* and *y* from independent normal distributions centered on the infector’s location. When an *x* or *y* located outside of the rectangular area of the simulated outbreak was sampled, we used a “mirror effect” to place it back into the rectangle using symmetry with the borders.

We drew the number of new mpox cases arising from spillover on day *t* from a Poisson distribution with mean equal to the daily spillover rate. The new case’s location was selected randomly within the 446 km x 413 km rectangle.

We ran the simulation for 5 years. For each simulation scenario, we performed 200 simulations. The reporting rate was assumed to be constant over time. For each simulation scenario, we used the same simulated epidemics but varied the reporting rate. To simulate different levels of reporting, we sampled observed cases from all cases according to each reporting probability considered in Table S8. The number of observed cases in the resulting datasets for each simulated scenario are shown in Figure S14.

We ran *vimes* on the simulated data and re-estimated *R_t_* and the spillover rate. When calculating the pairwise distances between cases, we calculated the temporal data as before, as the difference in days between symptom onset dates. For spatial data, we calculated Euclidean distances because the rectangular area used to simulate the data is flat.

Following Cori et al.,^14^ we used the true positive rate (TPR) and the true negative rate (TNR) to assess the ability of the model to correctly identify clusters of cases connected by transmission. The TPR, or sensitivity, is the proportion of case pairs which are connected by transmission that are inferred to be in the same cluster, while the TNR, or specificity, is the proportion of case pairs which are not connected by transmission that are inferred to not be in the same cluster. We compared the estimates of *R_t_* and the spillover rate with those used to simulate the epidemics by calculating the relative error.

We also used the results of the simulations to determine the optimal cutoff choice for mpox in Tshuapa Province. We defined the optimal cutoff as the one that had the highest mean of sensitivity and specificity across the 200 baseline simulations.

*Validation*. We searched ProMED for “monkeypox” on January 10, 2023. We searched Google Scholar for “monkeypox” AND “outbreak” AND “DRC” OR “Democratic Republic of Congo” on January 10, 2023.

**Supplementary results**

*Identifying clusters.* Using only temporal data, we identified one single large cluster of 934 cases (Figure S2). Using only spatial data, we identified 210 clusters, 62 of which were singletons. Using both data types combined, we obtained our main result consisting of 334 clusters (161 of which were singletons).

*Sensitivity analyses.* Results of the sensitivity analyses on the nine combinations of reporting rates and quantiles are shown in Figure S6 and Table S5. The length of the outbreaks (excluding singletons) is shown in Figure S7. In the most extreme scenario (98.3% quantile/10% reporting rate), the longest outbreak spanned 1,666 days. For our main result (98.3% quantile/25% reporting rate), the median outbreak length was 33 days (range 0 – 759).

Results using the shorter serial interval estimated by Miura et al.^15^ can be found in Figure S8 and Table S7. The main result using a quantile of 98.3% and reporting rate of 25% changed slightly: the estimated *R_t_* was 0.80 (95% CI: 0.79 – 0.85), and the annual spillover rate was 132 (95% CI: 122 – 143). As expected, outbreak lengths were shorter overall (Figure S9).

When we manually set the distance cutoff to 10 km, we obtained 212 clusters, of which 90 were singletons. The average cluster size was 4.4, and the maximum cluster size was 64. The estimated *R_t_* was 0.89 (95% CI: 0.86 – 0.93), and the annual spillover rate was 79 (72 – 87). Excluding singletons, mean outbreak length was 163 days (range 1 – 936).

When we performed the analysis separately for each year of data (using the year corresponding to symptom onset), mean cluster size increased from 2.3 in 2013 to 2.9 in 2017 (Table S2, Figure S10). Over the same period, the estimated *R_t_* increased slightly from 0.78 (95% confidence interval [CI]: 0.71 – 0.85) to 0.83 (95% CI: 0.76 – 0.90), and the annual spillover rate decreased from 199 (95% CI: 171 – 233) to 126 (95% CI: 106 – 152), assuming a reporting rate of 25% and a cutoff corresponding to the 98.3% quantile of the input distance distributions (Figure S11).

When we performed the analysis by health zone (using the same reporting rate and quantiles as above), mean cluster size ranged from 1.8 in Monkoto to 3.9 in both Busanga and Wema (Table S3, Figure S12). The estimated *R_t_* ranged from 0.69 (95% CI: 0.42 – 0.95) in Monkoto and 0.88 (95% CI: 0.80 – 0.96) and 0.88 (95% CI: 0.72 – 1.04) in Busanga and Wema respectively (Figure S13). Five health zones had an upper 95% CI for *R_t_* of at least 0.95. The annual spillover rate ranged from 3 (95% CI: 2 – 5) in Wema to 18 (95% CI: 15 – 22) in Djolu.

*Reconstruction results.* Except for the scenario where we used a cutoff corresponding to 50% of the input distance distributions and the scenario where we underestimated reporting, we obtained precise estimates for our reconstruction scenarios (low relative error, Figure S16).

*Documented mpox outbreaks*. In 2013, there were 104 possible mpox cases reported in Bokungu Health Zone, an increase that prompted an outbreak investigation. The results of this investigation were published in the literature. Of those, 60 were tested for MPXV: 50 cases tested positive, and 10 cases tested negative.^19^ Our analysis identified 11 mpox clusters in this health zone in 2013, 3 singletons, 3 clusters of size 2, 2 clusters of size 4, 1 cluster of size 6, and 2 clusters of size 8.

We identified one report of a possible mpox outbreak in Tshuapa Province from 2010 – 2019 on ProMED. According to the article, there were 20 suspected cases of mpox in the province in September 2015, with 18 cases being hospitalized in the city of Ikela in the Ikela Health Zone.^20^ We found only one cluster in our analysis that could have been part of this outbreak, a singleton with symptom onset date in June of that year.

*Mpox exposure history*. Out of 892 patients with available data, 418 (47%) reported contact with a person or persons presenting with similar symptoms in the three weeks prior to symptom onset; 346 (83%) of these 418 patients were assigned to clusters with size > 1 in our analysis (Figure S3). Information about the relationship was available for 377 out of 418 patients: 335 (89%) reported living with the ill contact and 154 (41%) reported sharing a bed with the ill contact. There were 829 patients with available data on animal exposures: 399 (48%) reported having touched a wild animal during three weeks prior to symptom onset; 322 (81%) of these 399 patients were assigned to clusters with size > 1. Patients reported exposure to a range of animals, with monkeys (253, 63%), rats (54, 14%), and squirrels (42, 11%) being the most common. Out of 800 patients with available data, 107 (13%) reported both animal and human exposures in the three weeks prior to symptom onset. Exposure history by health zone is shown in Figure S4.

**Supplementary figures**

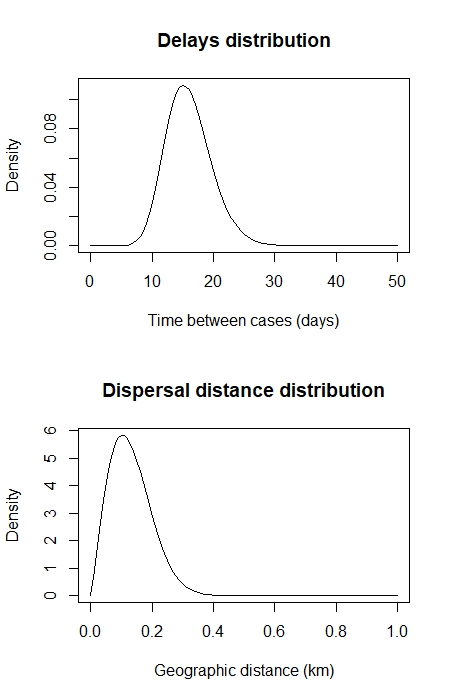

**Figure S1.** **Distributions of the serial interval (top) and distance kernel (bottom) used to identify clusters of human mpox cases in Tshuapa Province, DRC.**

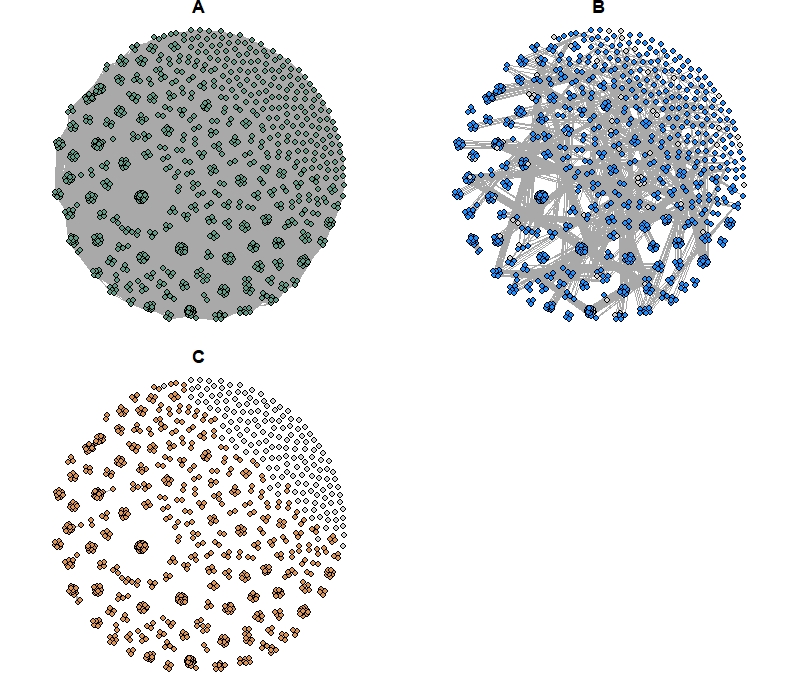

**Figure S2. Clusters of mpox cases in Tshuapa Province, DRC, 2013 – 2017 obtained using (A) only temporal distances, (B) only spatial distances, and (C) both data types combined.** Nodes represent cases, while edges represent possible epidemiological links. Cases in gray are not connected to other cases. We assumed a reporting rate of 25% and used pruning cutoff distances associated with the 98.3% quantiles of the input distance distributions.

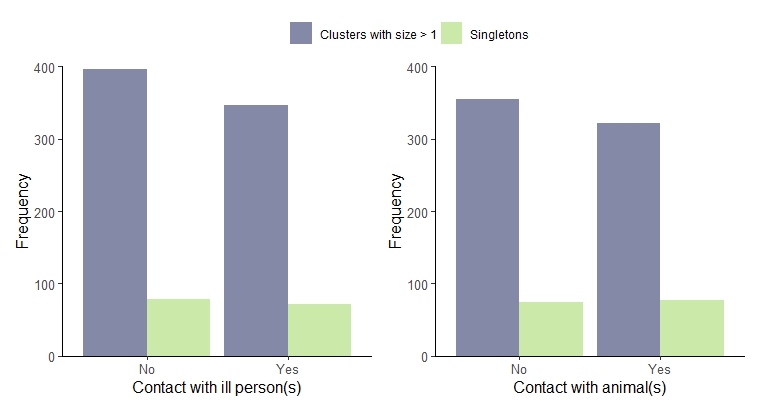

**Figure S3. MPXV exposure history by cluster type.** There were 892 patients with available data on contact with a person or persons presenting with similar symptoms in the three weeks prior to symptom onset and 829 patients with available data on animal exposures in the three weeks prior to symptom onset.

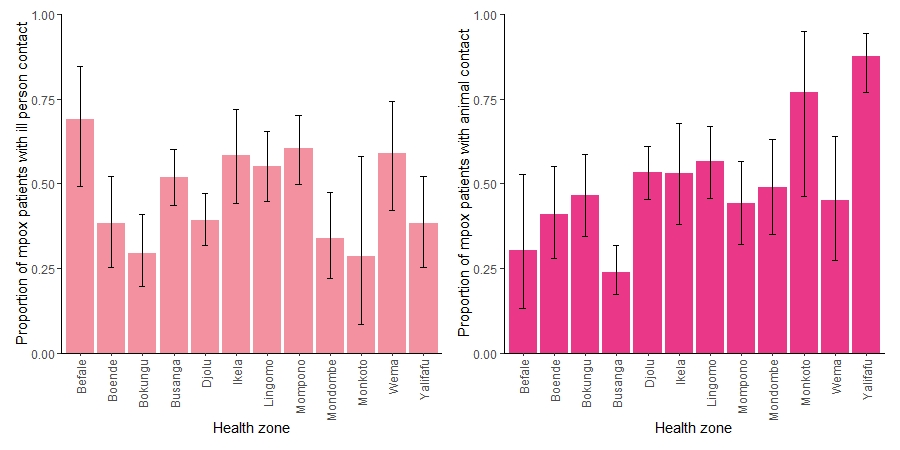

**Figure S4. MPXV exposure history by health zone.** There were 892 patients with available data on contact with a person or persons presenting with similar symptoms in the three weeks prior to symptom onset and 829 patients with available data on animal exposures in the three weeks prior to symptom onset. Error bars represent 95% exact binomial confidence intervals.

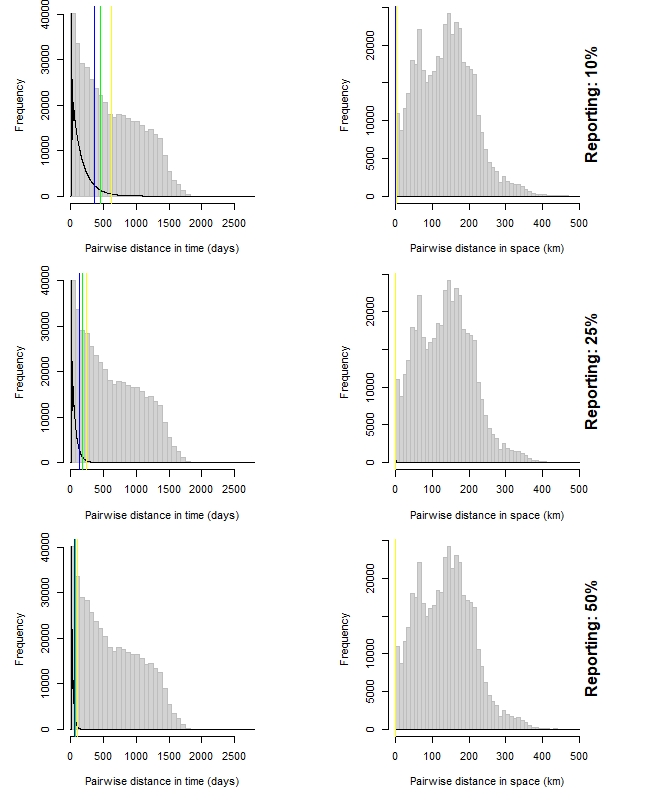

**Figure S5.** **Distributions of pairwise distances for reported human mpox cases in Tshuapa Province, DRC.** Temporal distance (left) is the time in days between symptom onset of the cases. Spatial distance (right) is the distance in km between the geographic locations of the cases calculated using the Vincenty inverse formula for ellipsoids. The observed pairwise distances between any two mpox cases is shown by the gray histograms. The input distribution of distances between a case and its closest observed ancestor assuming different reporting rates is shown by the black curved lines. The cutoffs associated with the 90%, 95%, and 98.3% quantiles of the distributions are indicated by the blue, green, and yellow vertical lines, respectively (only the yellow lines are visible on the spatial distance plots due to overplotting). Pairs of cases that have observed distances above the cutoff for each data type are deemed unconnected by transmission.

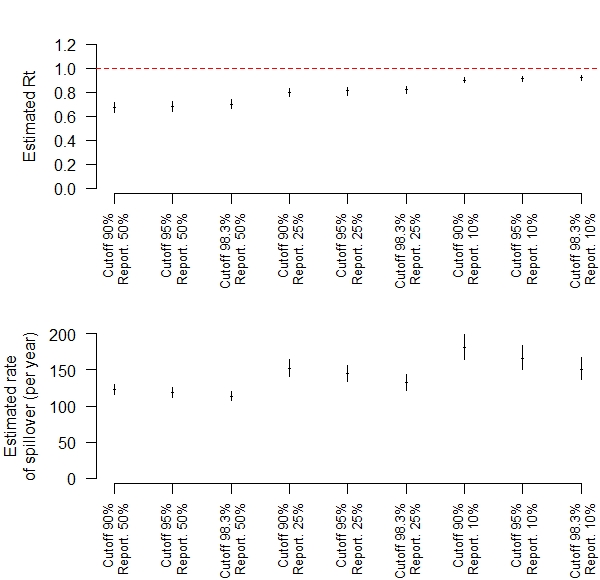

**Figure S6.** **Estimated effective reproduction number, *R_t_*, (top) and annual rate of spillover of monkeypox virus into the human population (bottom)** **in Tshuapa Province, DRC, 2013 – 2017 for different assumptions about the reporting rate (10%, 25%, or 50%) and cutoff for pruning (90%, 95%, or 98.3% quantile).** The maximum likelihood estimates are shown by the points, and the 95% confidence intervals are shown by the vertical bars. The threshold *R_t_* = 1 is shown by the horizontal dashed line in the top panel.

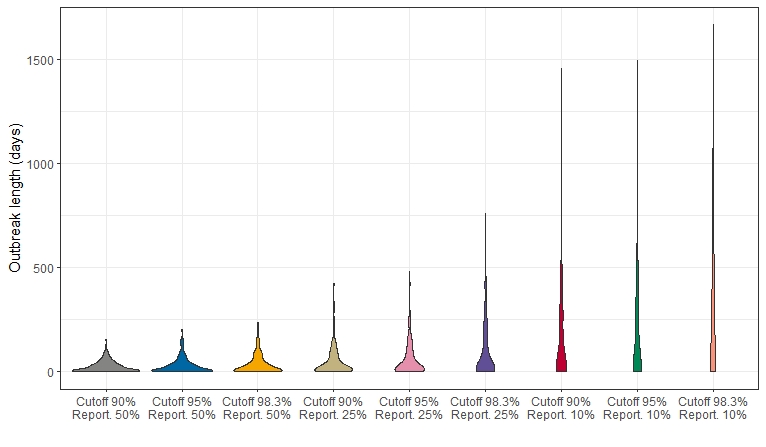

**Figure S7. Violin plots of the distribution of outbreak length in days in Tshuapa Province, DRC, 2013 – 2017 for different assumptions about the reporting rate (10%, 25%, or 50%) and cutoff for pruning (90%, 95%, or 98.3% quantile).** Only clusters with size > 1 were considered (singletons were excluded).

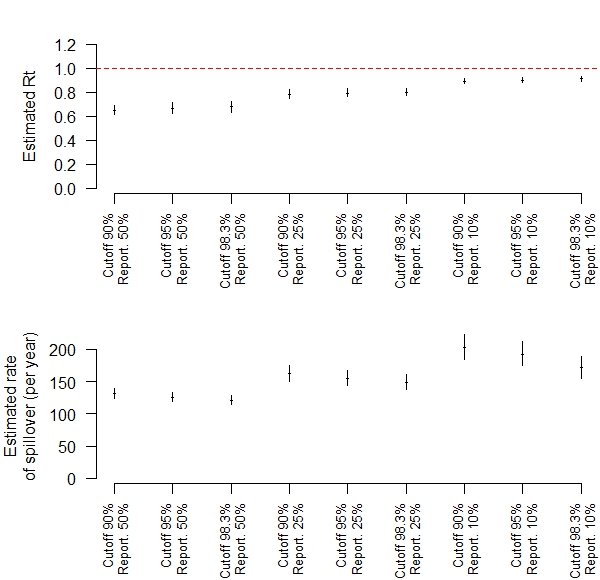

**Figure S8.** **Estimated effective reproduction number, *R_t_*, (top) and annual rate of spillover of monkeypox virus into the human population (bottom) in Tshuapa Province, DRC, 2013 – 2017 for different assumptions about the reporting rate (10%, 25%, or 50%) and cutoff for pruning (90%, 95%, or 98.3% quantile) using a serial interval with mean 10.3 days.** The maximum likelihood estimates are shown by the points, and the 95% confidence intervals are shown by the vertical bars. The threshold *R_t_* = 1 is shown by the horizontal dashed line in the top panel.

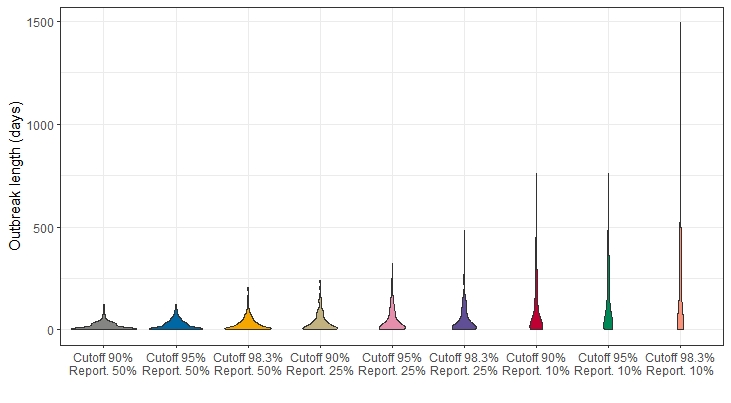

**Figure S9.** **Violin plots of the distribution of outbreak length in days in Tshuapa Province, DRC, 2013 – 2017 for different assumptions about the reporting rate (10%, 25%, or 50%) and cutoff for pruning (90%, 95%, or 98.3% quantile) using a serial interval with mean 10.3 days.** Only clusters with size > 1 were considered (singletons were excluded).

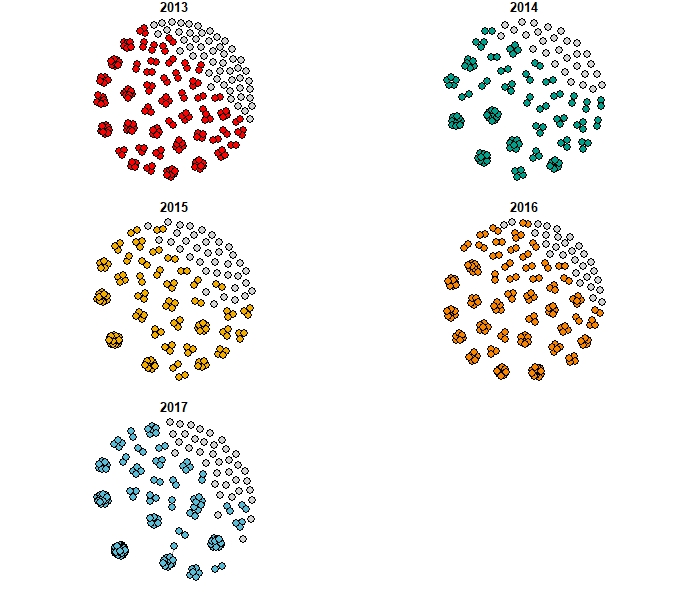

**Figure S10. Clusters of human mpox cases in Tshuapa Province, DRC by year 2013 – 2017.** We assumed a reporting rate of 25% and used pruning cutoff distances associated with the 98.3% quantiles of the input distance distributions. Cases in color were assigned to clusters, while cases in gray were not connected to other cases, using both temporal and spatial data.

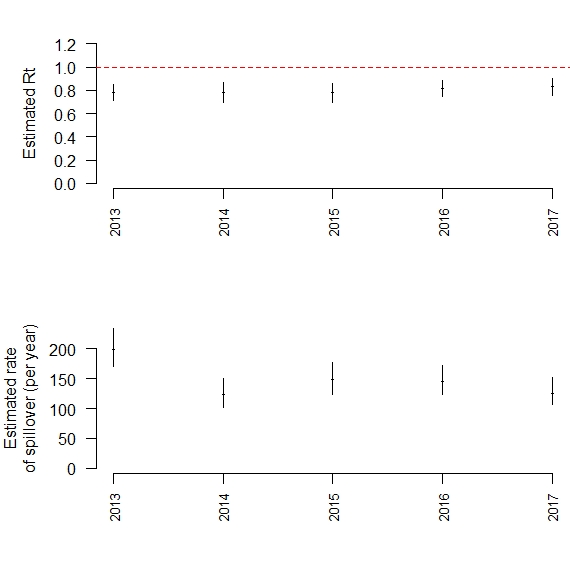

**Figure S11.** **Estimated effective reproduction number, *R_t_*, (top) and annual rate of spillover of monkeypox virus into the human population (bottom) in Tshuapa Province, DRC by year 2013 – 2017.** The maximum likelihood estimates are shown by the points, and the 95% confidence intervals are shown by the vertical bars. The threshold *R_t_* = 1 is shown by the horizontal dashed line in the top panel. We assumed a reporting rate of 25% and used pruning cutoff distances associated with the 98.3% quantiles of the input distance distributions.

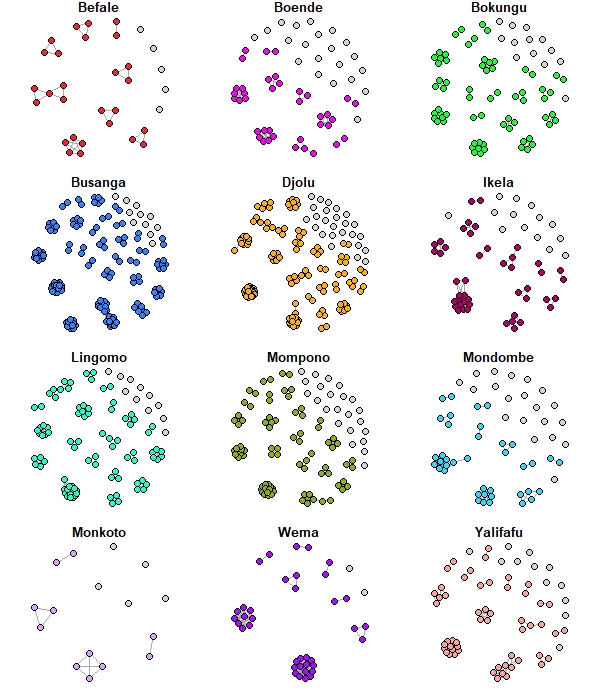

**Figure S12.** **Clusters of human mpox cases in Tshuapa Province, DRC by health zone, 2013 – 2017.** We assumed a reporting rate of 25% and used pruning cutoff distances associated with the 98.3% quantiles of the input distance distributions. Cases in color were assigned to clusters, while cases in gray were not connected to other cases, using both temporal and spatial data.

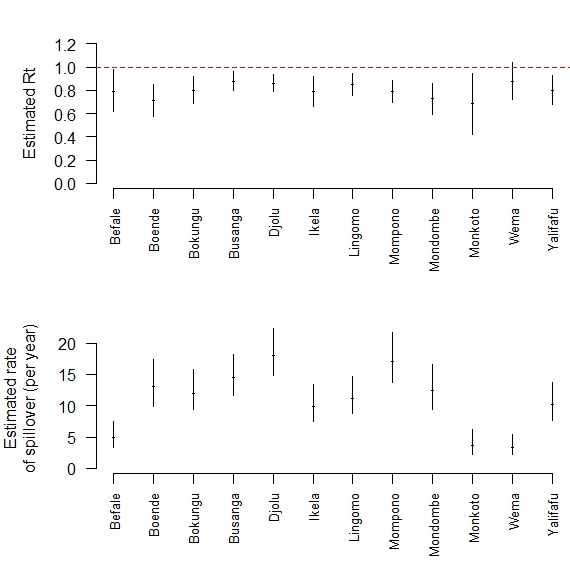

**Figure S13.** **Estimated effective reproduction number, *R_t_*, (top) and annual rate of spillover of monkeypox virus into the human population (bottom) in Tshuapa Province, DRC by health zone, 2013 – 2017.** The maximum likelihood estimates are shown by the points, and the 95% confidence intervals are shown by the vertical bars. The threshold *R_t_* = 1 is shown by the horizontal dashed line in the top panel. We assumed a reporting rate of 25% and used pruning cutoff distances associated with the 98.3% quantiles of the input distance distributions.

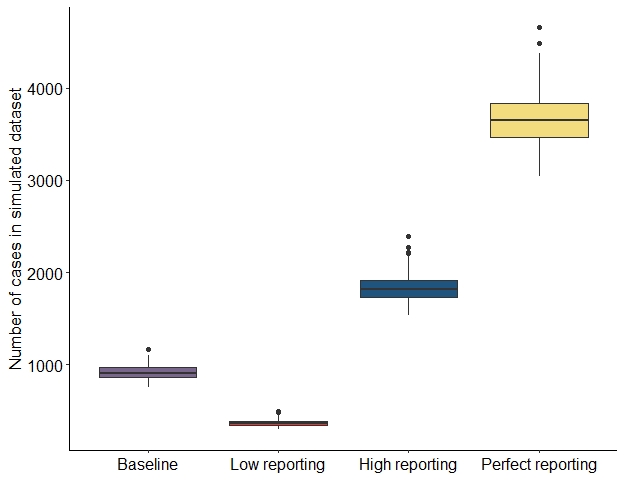

**Figure S14.** **Distribution of the simulated dataset sizes for each of the four simulation scenarios considered.**

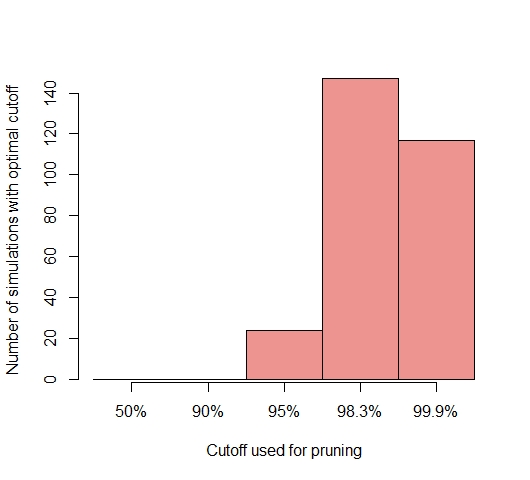

**Figure S15.** **Distribution of the optimal cutoff choice for mpox in Tshuapa Province, DRC.** The optimal cutoff had the highest mean of sensitivity and specificity across the 200 baseline simulations. Several cutoffs performed the same, and therefore, an individual simulation could contribute to more than one cutoff.

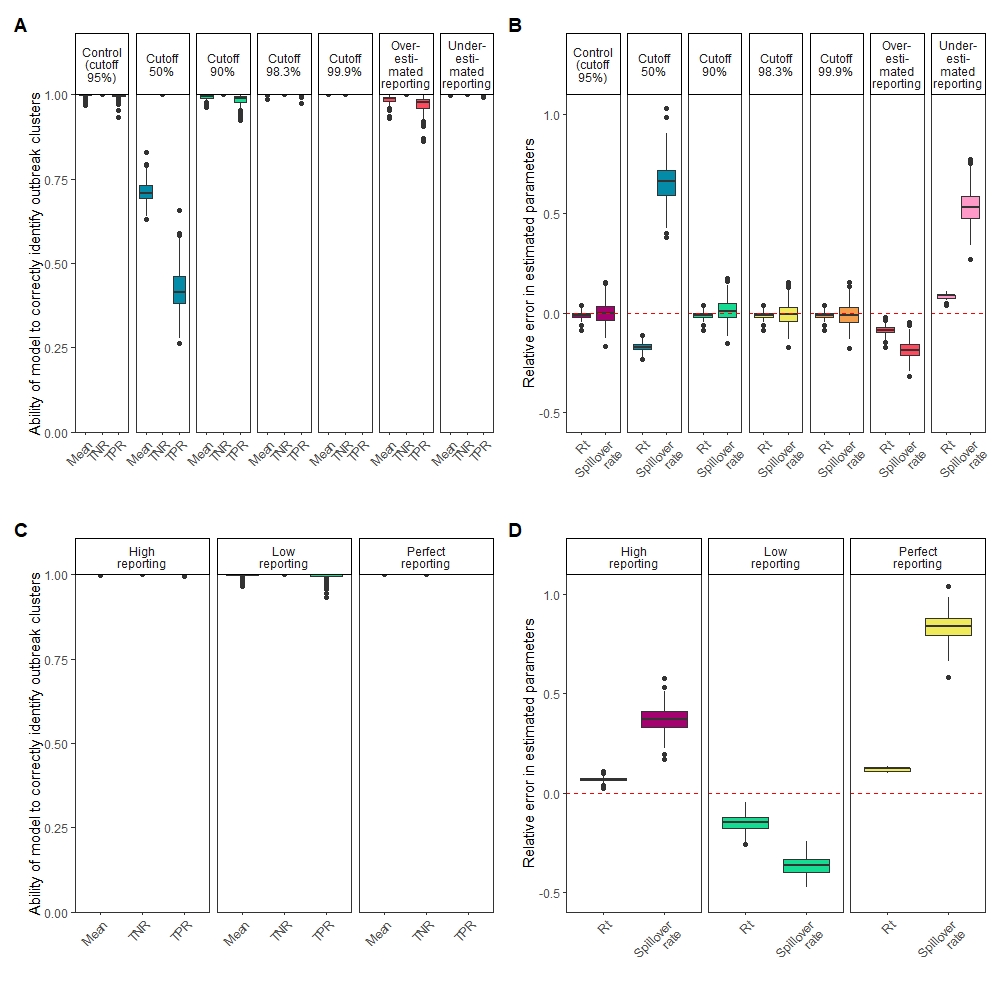

**Figure S16. The model performance for the control simulation using different reconstruction scenarios is shown in (A) and (B). The model performance for the baseline reconstruction scenario (using the optimal cutoff of 98.3%) applied to simulation scenarios with different reporting rates (low, high, and perfect) is shown in (C) and (D).** Model performance in (A) and (C) is shown in terms of the true positive rate (TPR), true negative rate (TNR), and the mean of the TPR and TNR. The relative error in the estimated reproduction number and spillover rate is shown in (B) and (D) across all scenarios.

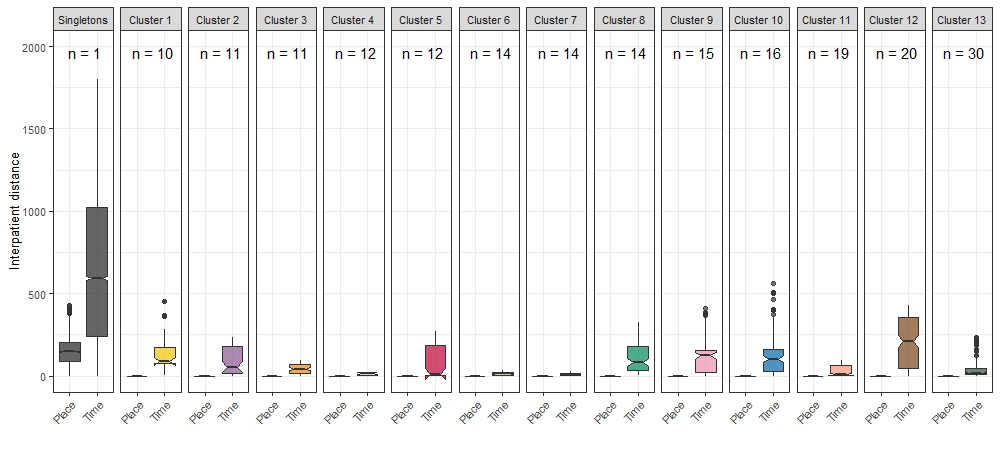

**Figure S17. Notched boxplots of the pairwise differences by data type and cluster of human mpox cases, Tshuapa Province, DRC, 2013 – 2017.** We assumed a reporting rate of 25% and used pruning cutoff distances associated with the 98.3% quantiles of the input distance distributions. There were 334 clusters, 161 of which were singletons. In addition to singletons, we plotted the clusters with 10 or more cases. The cluster size is indicated by “n” at the top of each panel. The interquartile range is shown by the boxes’ height, and the median of the pairwise differences is shown by the horizontal lines in the notches. The lower and upper bounds of the notches is considered the 95% tolerance interval. The height of the upper whisker is determined by the smaller of the 75^th^ percentile and the largest value, while the height of the lower whisker is determined by the larger of the 25^th^ percentile or the minimum value. Black points are possible outliers. The units for time are days, and the units for place are km. The most plausible clusters were clusters 3, 4, 6, 7, 11, and 13.

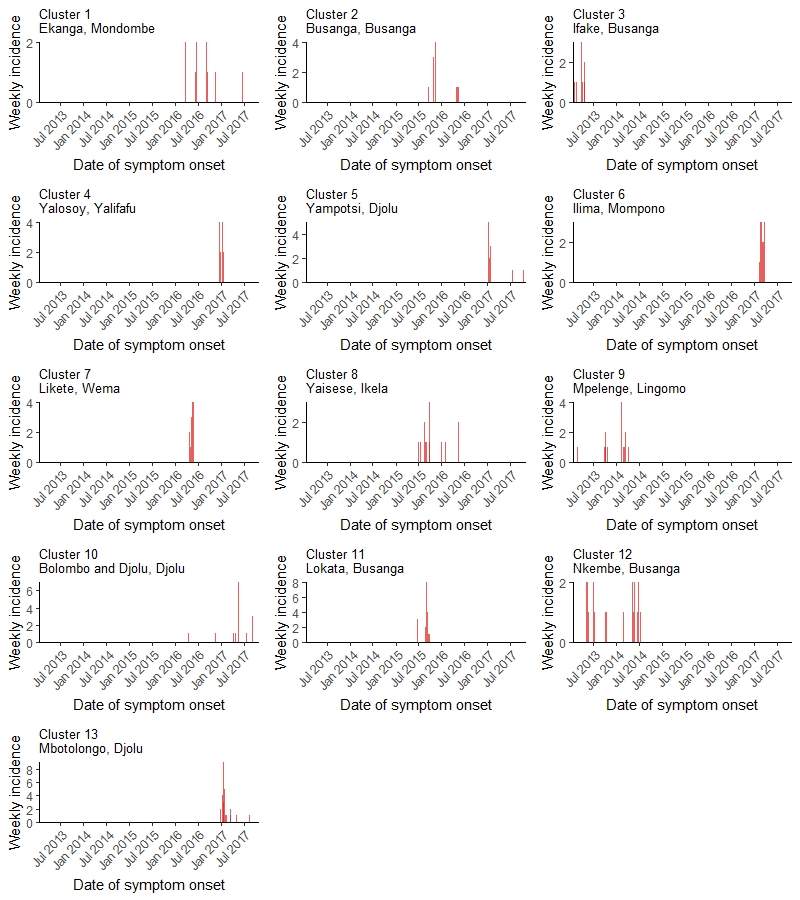

**Figure S18. Weekly incidence of human mpox cases by date of symptom onset for clusters with at least 10 cases, Tshuapa Province, DRC, 2013 – 2017.**

**Supplementary tables**

**Table S1. Published estimates of the basic reproduction number (*R_o_*) and effective reproduction number (*R_t_*) for mpox in DRC.**

| **Reference** | **Study year** | **Data years** | **Location** | **Type of reproduction number** | **Estimate (uncertainty)** |
| --- | --- | --- | --- | --- | --- |
| ^21^ | 1988 | 1980 – 1984 | DRC | *R_o_* | 0.82 (upper limit of 1) |
| ^22^ | 1988 | 1981 – 1986 | DRC | *R_t_* | 0.27** |
| ^23^ | 2005 | 1980 – 1984 | DRC | *R_t_* | 0.32 (0.22 – 0.40) |
| ^24^ | 2013 | 1980 – 1984 | DRC | *R_t_* | 0.30 (0.22 – 0.40) |
| ^25^* | 2015 | 2005 – 2007 | Sankuru District, DRC | *R_t_* | 0.58 |
| ^26^* | 2019 | 1982 – 1986 | DRC | *R_t_* | 0.38 (0.31 – 0.45) |
| ^27^ | 2020 | 1980 – 1984 | DRC | *R_o_* | 2.1 (1.5 – 2.7) |
| ^27^ | 2020 | 2011 – 2012 | DRC | *R_t_* | 0.85 (0.51 – 1.25) |
| This study | 2023 | 2013 – 2017 | Tshuapa Province, DRC | *R_t_* | 0.82 (0.79 – 0.85) |

*These studies have not been peer-reviewed.

**Calculated from the distribution of primary and presumed secondary cases by their generation rank (e.g., first generation, second generation, etc.) in Table 3 of Jezek et al.: (69/203 + 19/69 + 4/19 + 1/4)/4 = 0.27. The study authors assumed a rash serial interval of 14 days when determining generation rank.

**Table S2.** **Characteristics of clusters of human mpox cases in Tshuapa Province, DRC by year, 2013 – 2017. Clusters were identified using an algorithm implemented in the R package *vimes*.**

| **Year** | **Number of clusters** | **Number of singletons** | **Maximum cluster size** | **Mean cluster size** |
| --- | --- | --- | --- | --- |
| 2013 | 98 | 51 | 11 | 2.3 |
| 2014 | 61 | 29 | 10 | 2.3 |
| 2015 | 73 | 37 | 19 | 2.3 |
| 2016 | 74 | 29 | 14 | 2.8 |
| 2017 | 65 | 37 | 28 | 2.9 |

**Table S3.** **Characteristics of clusters of human mpox cases in Tshuapa Province, DRC by health zone, 2013 – 2017.** Clusters were identified using an algorithm implemented in the R package *vimes*.

| **Health zone** | **Number of clusters** | **Number of singletons** | **Maximum cluster size** | **Mean cluster size** |
| --- | --- | --- | --- | --- |
| Befale | 13 | 5 | 5 | 2.5 |
| Boende | 31 | 21 | 7 | 1.9 |
| Bokungu | 31 | 13 | 8 | 2.6 |
| Busanga | 39 | 11 | 20 | 3.9 |
| Djolu | 48 | 25 | 30 | 3.5 |
| Ikela | 25 | 14 | 14 | 2.4 |
| Lingomo | 30 | 11 | 15 | 3.3 |
| Mompono | 43 | 23 | 14 | 2.5 |
| Mondombe | 30 | 20 | 10 | 2.0 |
| Monkoto | 9 | 5 | 4 | 1.8 |
| Wema | 10 | 2 | 14 | 3.9 |
| Yalifafu | 26 | 12 | 12 | 2.5 |

**Table S4.** **Cutoffs for pairwise distances considered in sensitivity analyses.**

|  |  | **Cutoff 90%** | **Cutoff 95%** | **Cutoff 98.3%** |
| --- | --- | --- | --- | --- |
| **Temporal (days)** | **Reporting 50%** | 62 | 78 | 103 |
|  | **Reporting 25%** | 136 | 175 | 233 |
|  | **Reporting 10%** | 356 | 460 | 619 |
| **Spatial (km)** | **Reporting 50%** | 0.3 | 0.4 | 0.5 |
|  | **Reporting 25%** | 0.5 | 0.6 | 0.7 |
|  | **Reporting 10%** | 0.7 | 0.9 | 1.2 |

**Table S5. Sensitivity analysis on quantiles and reporting rates for identifying clusters of human mpox cases in Tshuapa Province, 2013 – 2017.**

|  | **Number of clusters** | **Number of singletons** | **Maximum cluster size** | **Mean cluster size** |
| --- | --- | --- | --- | --- |
| **Cutoff 90%**  **Reporting 50%** | 420 | 231 | 29 | 2.2 |
| **Cutoff 95%**  **Reporting 50%** | 407 | 221 | 29 | 2.3 |
| **Cutoff 98.3%**  **Reporting 50%** | 392 | 209 | 29 | 2.4 |
| **Cutoff 90%**  **Reporting 25%** | 376 | 194 | 30 | 2.5 |
| **Cutoff 95%**  **Reporting 25%** | 361 | 184 | 30 | 2.6 |
| **Cutoff 98.3%**  **Reporting 25%** | 334 | 161 | 30 | 2.8 |
| **Cutoff 90%**  **Reporting 10%** | 303 | 138 | 30 | 3.1 |
| **Cutoff 95%**  **Reporting 10%** | 282 | 123 | 30 | 3.3 |
| **Cutoff 98.3%**  **Reporting 10%** | 260 | 100 | 30 | 3.6 |

**Table S6. Cutoffs for pairwise distances considered in sensitivity analyses using a serial interval with mean 10.3 days.**

|  |  | **Cutoff 90%** | **Cutoff 95%** | **Cutoff 98.3%** |
| --- | --- | --- | --- | --- |
| **Temporal (days)** | **Reporting 50%** | 43 | 54 | 72 |
|  | **Reporting 25%** | 90 | 116 | 156 |
|  | **Reporting 10%** | 232 | 300 | 404 |
| **Spatial (km)** | **Reporting 50%** | 0.3 | 0.4 | 0.5 |
|  | **Reporting 25%** | 0.5 | 0.6 | 0.7 |
|  | **Reporting 10%** | 0.7 | 0.9 | 1.2 |

**Table S7. Sensitivity analysis on quantiles and reporting rates for identifying clusters of human mpox cases in Tshuapa Province, 2013 – 2017 using a serial interval with mean 10.3 days.**

|  | **Number of clusters** | **Number of singletons** | **Maximum cluster size** | **Mean cluster size** |
| --- | --- | --- | --- | --- |
| **Cutoff 90%**  **Reporting 50%** | 445 | 255 | 29 | 2.1 |
| **Cutoff 95%**  **Reporting 50%** | 428 | 237 | 29 | 2.2 |
| **Cutoff 98.3%**  **Reporting 50%** | 415 | 228 | 29 | 2.3 |
| **Cutoff 90%**  **Reporting 25%** | 396 | 211 | 29 | 2.4 |
| **Cutoff 95%**  **Reporting 25%** | 381 | 199 | 30 | 2.5 |
| **Cutoff 98.3%**  **Reporting 25%** | 369 | 189 | 30 | 2.5 |
| **Cutoff 90%**  **Reporting 10%** | 334 | 161 | 30 | 2.8 |
| **Cutoff 95%**  **Reporting 10%** | 319 | 148 | 30 | 2.9 |
| **Cutoff 98.3%**  **Reporting 10%** | 290 | 128 | 30 | 3.2 |

**Table S8. Simulation scenarios.**

| **Simulation scenario** | **Reproduction number** | **Mean serial interval in days (standard deviation)** | **Standard deviation of spatial kernel** | **Spillover rate (introductions per year)** | **Reporting rate** |
| --- | --- | --- | --- | --- | --- |
| **Baseline** | 0.81 (our estimate for mpox) | 16.0 (3.7)^9^ | 0.10 km (estimated from WHO contact tracing data) | 145 (our estimate) | 25% (adjusted estimate from Nolen et al.^19^ for missing data) |
| **Low reporting** |  |  |  |  | 10% |
| **High reporting** |  |  |  |  | 50% |
| **Perfect reporting** |  |  |  |  | 100% |

**Table S9. Reconstruction scenarios.**

| **Reconstruction scenario** | **Quantile used for cutoff** | **Reporting rate** |
| --- | --- | --- |
| Control | 0.95 | Same as simulation |
| 50% cutoffs | 0.50 | Same as simulation |
| 90% cutoffs | 0.90 | Same as simulation |
| 98.3% cutoffs | 0.983 | Same as simulation |
| 99.9% cutoffs | 0.999 | Same as simulation |
| Underestimated reporting | 0.95 | 0.1 |
| Overestimated reporting | 0.95 | 0.4 |

3.	OpenStreetMap RDC, Democratic Republic of Congo (DRC) Localities (OpenStreetMap Export). Available at: <https://data.humdata.org/dataset/democratic-republic-of-congo-drc-localities-openstreetmap-export>. Accessed.

4.	American Red Cross, DRC Health Zone and Health Area boundaries. Available at: <https://data.humdata.org/m/dataset/drc-health-data>? Accessed January 9, 2023.

5.	UCLA DRC, 2010. DRC localities. RGC, ed. Map for Environment.

6.	National Geospatial-Intelligence Agency, Geographic Names Server. Available at: <https://geonames.nga.mil/geonames/GNSHome/index.html>. Accessed.

7.	Google, Google Earth. Available at: <https://earth.google.com/web/>. Accessed.

8.	van der Loo M, 2014. The stringdist Package for Approximate String Matching. The R Journal 6: 111-122.

9.	Nishiura H, Eichner M, 2007. Infectiousness of smallpox relative to disease age: estimates based on transmission network and incubation period. Epidemiology & Infection 135: 1145-1150.

10.	Rohatgi A, 2021. WebPlotDigitizer. Pacifica, California, USA.

11.	Fenner F, Henderson D, Arita I, Jezek Z, Ladnyi I, 1988. Smallpox and its Eradication: World Health Organization.

12.	Jezek Z, Fenner F, 1988. Human Monkeypox (Monographs in Virology). Basel, Switzerland: Karger.

13.	Wallace J, 2010. R Package 'Imap'. CRAN.

14.	Cori A, Nouvellet P, Garske T, Bourhy H, Nakouné E, Jombart T, 2018. A graph-based evidence synthesis approach to detecting outbreak clusters: An application to dog rabies. PLoS Comput Biol 14: e1006554.

15.	Miura F, Backer J, van Rijckevorsel G, Bavalia R, Raven S, Petrignani M, Ainslie K, Wallinga J, Dutch Mpox Response Team, 2022. Time scales of human mpox transmission in the Netherlands. medRxiv.

16.	Jombart T, 2017. outbreaker: GitHub.

17.	Jombart T, 2018. quicksim: GitHub.

18.	O'Driscoll M, Harry C, Donnelly C, Cori A, Dorigatti I, 2021. A comparative analysis of statistical methods to estimate the reproduction number in emerging epidemics, with implications for the current coronavirus disease 2019 (COVID-19) pandemic. Clinical Infectious Diseases 73: e215-e223.

19.	Nolen L, Osadebe L, Katomba J, Likofata J, Mukadi D, Monroe B, Doty J, Hughes C, Kabamba J, Malekani J, 2016. Extended Human-to-Human Transmission during a Monkeypox Outbreak in the Democratic Republic of the Congo. Emerg Infec Dis 22: 1014-21.

20.	Herriman R, 2015. Suspected monkeypox outbreak reported in DRC. Outbreak News Today.

21.	Fine P, Jezek Z, Grab B, Dixon H, 1988. The Transmission Potential of Monkeypox Virus in Human Populations. International Journal of Epidemiology 17: 643-650.

22.	Jezek Z, Grab B, Szczeniowski M, Paluku K, Mutombo M, 1988. Human monkeypox: secondary attack rates. Bull World Health Organ 66: 465-470.

23.	Lloyd-Smith J, Schreiber S, Kopp P, Getz W, 2005. Superspreading and the effect of individual variation on disease emergence. Nature 428: 355-359.

24.	Blumberg S, Lloyd-Smith J, 2013. Inference of R0 and Transmission Heterogeneity from the Size Distribution of Stuttering Chains. PLoS Comput Biol 9: e1002993.

25.	McMullen C, 2015. A One Health Perspective on Disease Dynamics: Human Monkeypox Transmission in Sankuru District, Democratic Republic of Congo. Duke Global Health Institute: Duke University.

26.	Ambrose M, Kucharski A, Formenty P, Muyembe-Tamfum J, Rimoin A, Lloyd-Smith J, 2019. Quantifying transmission of emerging zoonoses: Using mathematical models to maximize the value of surveillance data. bioRxiv.

27.	Grant R, Nguyen LL, Breban R, 2020. Modelling human-to-human transmission of monkeypox. Bull World Health Organ 98: 638-640.
